# COVID-19 in China: Risk Factors and R_0_ Revisited

**DOI:** 10.1101/2020.05.18.20104703

**Authors:** Irtesam Mahmud Khan, Wenyi Zhang, Sumaira Zafar, Yong Wang, Junyu He, Hailong Sun, Ubydul Haque, M Sohel Rahman

## Abstract

The COVID-19 epidemic had spread rapidly through China and subsequently has proliferated globally leading to a pandemic situation around the globe. Human-to-human transmissions, as well as asymptomatic transmissions of the infection, have been confirmed. As of April 3^rd^’ public health crisis in China due to COVID-19 is potentially under control. We compiled a daily dataset of case counts, mortality, recovery, temperature, population density, and demographic information for each prefecture during the period of January 11 to April 07, 2020 (excluding Wuhan from our analysis due to missing data). Understanding the characteristics of spatiotemporal clustering of the COVID-19 epidemic and R_0_ is critical in effectively preventing and controlling the ongoing global pandemic. The prefectures were grouped based on several relevant features using unsupervised machine learning techniques. We performed a computational analysis utilizing the reported cases in China to estimate the revised R_0_ among different regions for prevention planning in an ongoing global pandemic. Finally, our results indicate that the impact of temperature and demographic (different age group percentage compared to the total population) factors on virus transmission may be characterized using a stochastic transmission model. Such predictions will help prioritize segments of a given community/region for action and provide a visual aid in designing prevention strategies for a specific geographic region. Furthermore, revised estimation and our methodology will aid in improving the human health consequences of COVID-19 elsewhere.

## INTRODUCTION

The world is facing an unprecedented pandemic of a virus (COVID-19) that originated in Wuhan, China. As such, the virus has been declared a global public health emergency by the World Health Organization (WHO). The virus is of the genus *betacoronavirus* which are zoonotically transmitted enveloped RNA viruses (*1*). Certain strains of Coronavirus are known to cause infections in humans and have led to previous epidemics, such as severe acute respiratory syndrome (SARS) and Middle East respiratory syndrome (MERS) (*2*). A new strain of Coronavirus called the COVID-19 has led to a global epidemic since its emergence in Wuhan, China (*3*). As of May 17, 2020, this disruptive pandemic has been spread among 180 countries and infected >4.6 million individuals and caused >0.31 million deaths since the onset of the epidemic. Nearly 0.084 million cases were diagnosed in mainland China (*4*, *5*). The epidemic in China is now under control.

Initial efforts to stall transmission of the COVID-19 by the Chinese authorities included the closure of the Huanan Seafood Wholesale Market which was considered to be the focal point of the outbreak (*6*). However, in spite of such strong intervention attempts, the rapid spread of the virus could not be prevented. Apart from measures to prevent exposure to the source, diagnostic recognition of the virus is an essential part of preventing efforts to prevent transmission. Diagnostic tests for COVID-19 were developed after isolation of the virus from lower respiratory tract specimens and blood serum (*7*). Scientists in China provided the genetic sequence of COVID-19 to the WHO, which has in turn, made Real-Time Polymerase Chain Reaction (RT-PCR) tests available globally.(*7*) COVID-19 has a clinically milder presentation and lower case fatality ratio (CFR) when compared to SARS and MERS (*8*) and hence, there seems to be an increased risk of asymptomatic or pre-clinically diagnosed individuals spreading the disease (*9*).

With the current state of globalization and increased interconnectedness of the world, the risk of infectious disease spread has increased. A revised estimation of R_0_ based on some potential risk factors is one of the powerful tools for assessing an epidemic’s ability to spread (*10, 11*). They provide vital information regarding where resources for containment and treatment should be targeted. Additionally, the case definition of COVID-19 has been updated by the Chinese authorities to now include those who present clinically with evidence of radiographic changes on CT scans of the lungs. Infection rate and mortality are higher among male compare with female (*12*). The mortality rate is higher among the older age groups (*12*). Population density might be one of the potential risk factors in the spread of COVID-19 infections (*13*). Control effort proved one of the keys to prevent and contain the spatial spread (*14*). The role of temperature in the spread of COVID-19 has been studied (*15-22*) and debated among policymakers.

A revised estimation based on the geographic distribution of population density, age group, gender, temperature, morbidity, mortality, and recovery rate from COVID-19 on a contained epidemic country is necessary in order to accurately predict the spatial spread of the virus and identify the risk factors for targeted control. Our study models the spread of COVID-19 among three regions in China, where the regions are not clustered geographically; rather they are intelligently grouped or clustered based on several relevant features (mentioned above) through application of unsupervised machine learning techniques. Thus, it will provide more accurate findings regarding where public health measures should be targeted.

## MATERIALS AND METHODS

### Data Source

In this study, 31 provinces of Mainland China were taken into consideration, including 367 county-level administrative areas. The number of daily confirmed, cured, and death of COVID-19 cases were collected from the National Health Commission of the People’s Republic of China (http://www.nhc.gov.cn/) and the provincial health commissions.

COVID-19 that started to spread from Wuhan in the mainland of China is a notifiable disease (*23*). Local health professionals (responsible for investigation and exposure information collection) tied to add the daily reported cases in China’s Infectious Disease Information System (IDIS). IDIS system recorded all cases with identification numbers, to avoid data duplication. All data contained information about morbidity, mortality, and recovery from COVID-19 case records in the IDIS through the end of April 3, 2020, were extracted. IDIS system categorized all the confirmed cases as suspected, clinically diagnosed (Hubei Province only), or asymptomatic. Confirmed cases were diagnosed by testing the viral nucleic acid in throat swabs specimens (some samples tested retrospectively) (*23*). Suspected cases were diagnosed based on the clinical symptoms and exposure and were classified as clinically diagnosed cases. Clinically diagnosed cases include suspicious cases with lung imaging features consistent with coronavirus pneumonia. The positive viral nucleic acid test is used to diagnose asymptomatic cases (without any COVID19 symptoms, e.g., fever, dry cough) (*23*). The date of diagnosis, recovery, and mortality were used as an onset date of infection in the time series data (*23*).

### Population

Demographic data for each city were obtained from the National Bureau of Statistics of China (http://www.stats.gov.cn/).

### Temperature

The daily surface air temperature dataset from a joint project of National Centres for Environmental Prediction (NCEP) and the National Center for Atmospheric Research (NCAR) is obtained at the prefecture-level of China. The NCEP/NCAR Reanalysis dataset is continuously available (1948 - present) at the global gridded dataset of earth atmosphere, incorporating the in-situ observations and numerical weather prediction (NWP) model output.

The temporal resolution is 6-hour (0000, 0600, 1200, and 1800 UTC) with a spatial resolution of 2.5-degree.

Google earth engine platform was used to download the required data following a three-step process: selection of required data, extraction to geographical location, and export to google drive. Data selection parameters include, dataset identifier (NCEP_RE/surface_temp), geographical location (China prefecture level), and date (2019-12-01 – 2020-04-07). To get the temperature data for each prefecture, ee.Reducer.first() function is used to get the first (or only) value of temperature. Finally, the extracted dataset is exported to the google drive as a CSV file. The temperature dataset consists of four observations per day and is used to obtain minimum, maximum, and mean temperatures.

### Computational Analysis

We divided China into three different regions applying a carefully designed clustering exercise as described below. The full analysis pipeline is illustrated in Figure 1 Supplement. Initially, all the prefectures that reported zero cases were excluded from the analysis. Then we applied an unsupervised clustering algorithm, namely, K-means algorithm, based on several features (Table 1, Supplement) and identified three separate regions within mainland China. Now, different sets of features (i.e., criteria used for clustering) may result in different outputs (i.e., different regional grouping). To analyse this, we applied our clustering algorithm on four different sets of features/criteria that include incidence, recovery rate, mortality rate, male-female ratio, age group ratio, minimum, mean and maximum temperature in each prefecture. Table 1 lists the criteria/features used for clustering (further details are provided in Table 1, Supplement). For visualization purposes, Principal Component Analysis (PCA) has been performed to reduce the dimensions of the feature set to two dimensions (Figure 2 and 3, supplement).

**Table 1:** Five different sets of criteria/features for clustering. The features used are, Incidence (I), Recovery Rate (RR), Mortality Rate (MR), Male/Female Ratio (MFR), Age Group Ratio (AGR), and Temperature (T). More details can be found Table 1 (Supplement).

| Cluster | Feature Set |
| --- | --- |
| A | I, RR, MR, MFR, AGR<sub>>64</sub>, T<sub>min</sub>, T<sub>avg</sub>, T<sub>max</sub> |
| B | I, RR, MR, AGR<sub>>64</sub>, T<sub>min</sub>, T<sub>avg</sub>, T<sub>max</sub> |
| C | I, RR, MR, AGR<sub><15</sub>, AGR<sub>15-64</sub>, AGR<sub>>64</sub>, T<sub>min</sub>, T<sub>avg</sub>, T<sub>max</sub> |
| D | T<sub>min</sub>, T<sub>avg</sub>, T<sub>max</sub> |

**Fig. 1.**
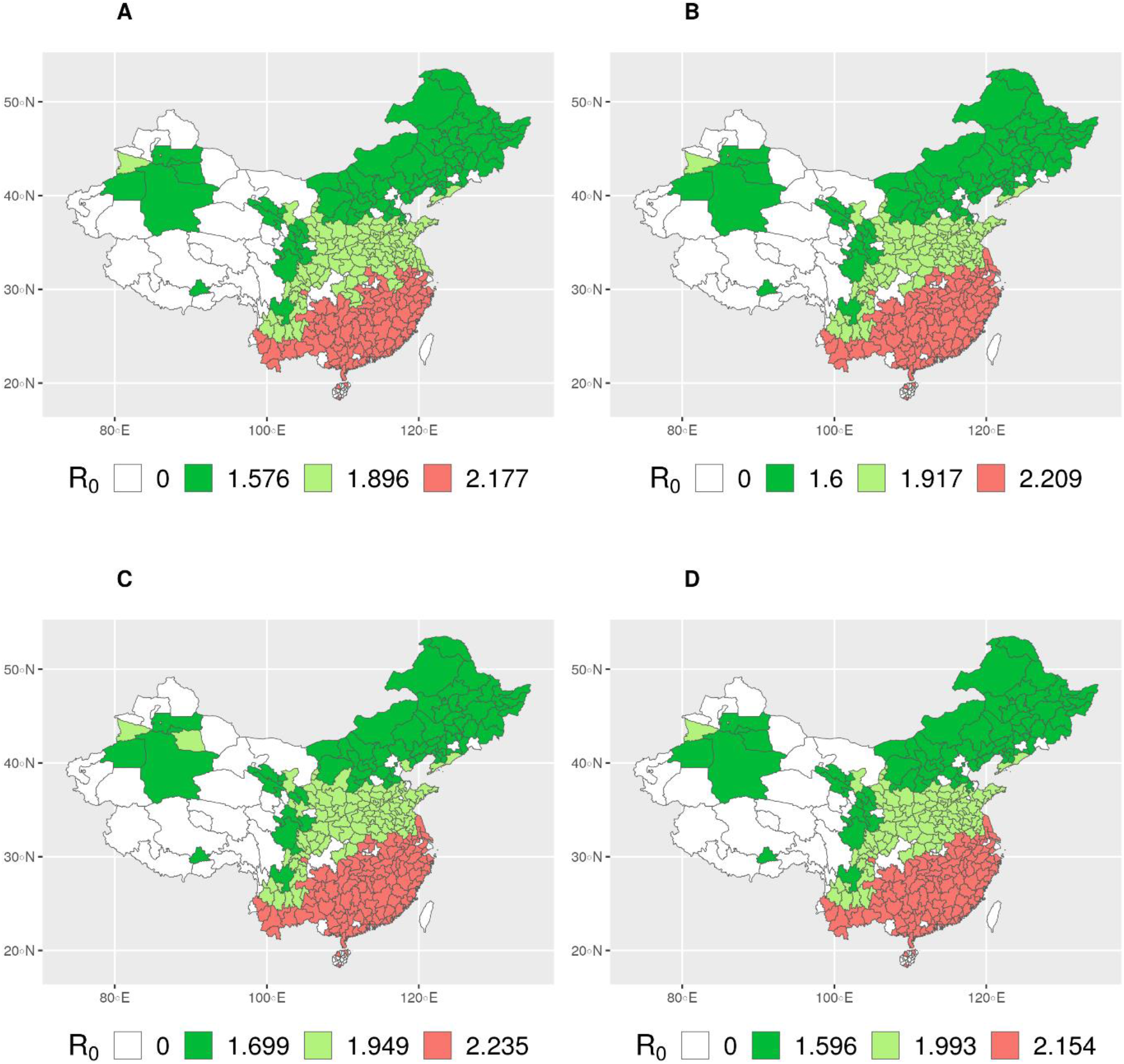
R_0_ values for different regions in China. A, B, C, and D indicates the clustering based on different sets of features. R0 value of 0 indicates no cases in that prefecture.

**Fig. 2.**
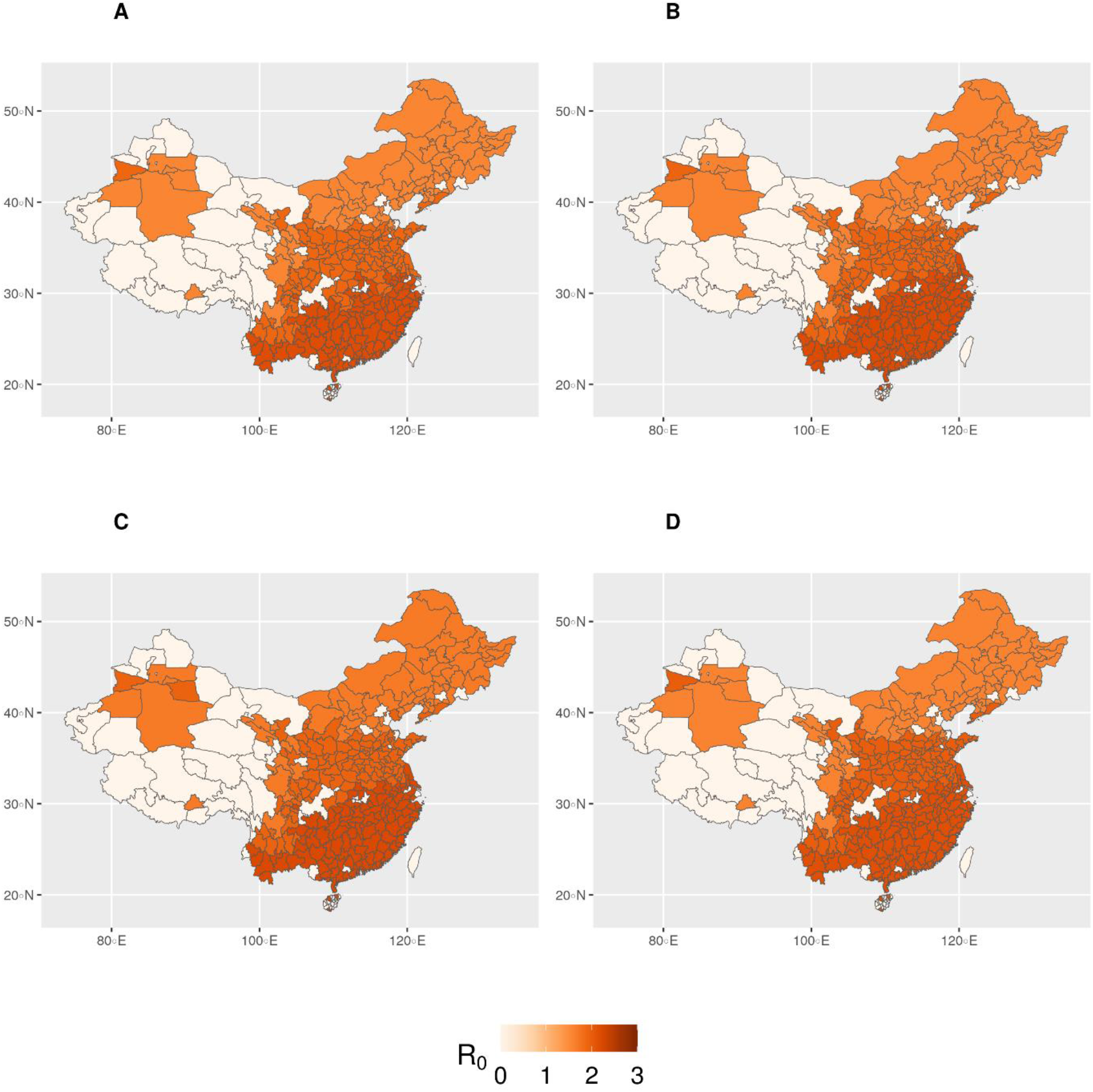
R_0_ values for different regions in China (scaled to 0-3). A, B, C, D indicates the clustering based on different sets of features. White colour indicates no cases in that prefecture. Darker shade of red corresponds to higher R_0_ and vice versa.

**Fig. 3.**
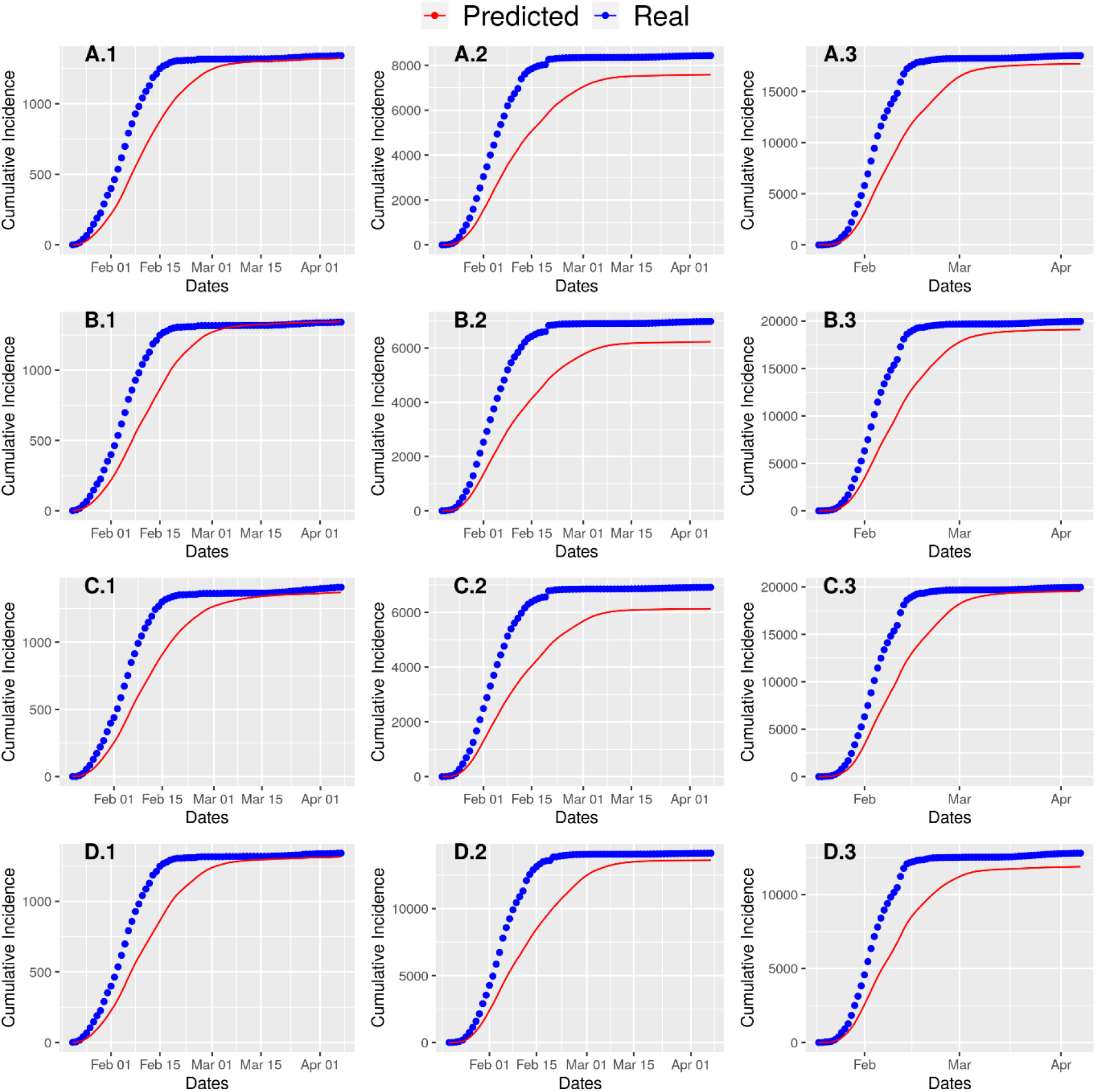
Real cumulative cases of COVID-19 and predicted cumulative cases by our model are plotted together. A.X corresponds to Region X of Clustering scheme A.

Incidence, mortality rate, and recovery rate is calculated as follows:

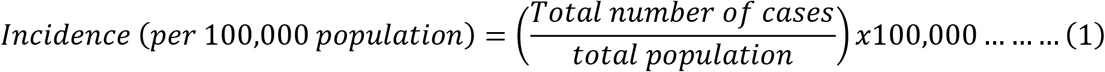

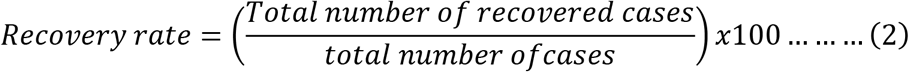

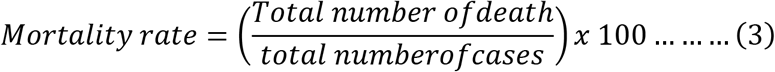

The K-means clustering algorithm (with default parameters) was used to do the clustering (*24*) (*25, 26*). In our dataset, there are 302 prefectures with at least one incidence. However, we have population and temperature data was available, for 298 and 296 prefectures, respectively. That is why 298 prefectures were considered for clustering B. On the other hand, 296 prefectures were considered for all other clusters. Notably, Wuhan was excluded from the analysis, because data for early days of transmission for Wuhan is not available (*27*).

After dividing all the prefectures into three different regions, a stochastic transmission dynamic model to estimate R_0_ for each region was fit. The daily incidence, recovery, and mortality of each region were fitted in the model. For estimating R_0_ of a region, we considered the daily confirmed incidences for all the prefectures of that region. The same procedure is applied for recovery and mortality.

We divided the population into three different compartments, namely, susceptible, infected and removed (SIR model (*28*), i.e., isolated, recovered, dead, or otherwise no longer infectious). The rate of change from susceptible to infected is termed as the rate of transmission. Similarly, the rate of change from infected to removed is termed as the removal rate. Following the work of Kucharski et al. (*29*), in our model, we have assumed that the delay distribution from the onset to isolation follows the well-known Erlang distribution with a mean of 2.9 days and a standard deviation of 2.1 days (*30*). So the removal rate is assumed to be 0.34 (1/2.9) (*31, 32*). Transmission is modelled as a stochastic random walk process and sequential Monte Carlo simulation is used to infer the transmission rate over time, resulting number of cases, and the time varying basic reproduction number (R_t_). Sequential Monte Carlo (i.e., particle filter) simulation (*29*) is run 100 times with bootstrap fits. R_0_ is then estimated by taking the median of the first 14 days of **R_t_**. For fitting time series incidence, mortality, and recovery data, we tried to maximize negative log-likelihood.

### Model Validation

The primary validation of the model is done through visually inspecting the plot generated using the model-inferred a number of cases and the actual cases. Then we calculated the Root Mean Square Error (RMSE) values from the cumulative real and predicted incidences (Table 2, supplement). While calculating the RMSE value for a particular cluster (under a particular clustering scheme), we compared the cumulative true number of cases against the predicted one for each day.

**Table 2:** Different regional results for different clustering schemes are reported in this table. The ranges within brackets refer to 95% Confidence Interval (CI) of R_0_.

|  |  | **A** | **B** | **C** | **D** |
| --- | --- | --- | --- | --- | --- |
| Regions | **1** | 1.58 (1.25-1.99) | 1.60 (1.30-1.93) | 1.70 (1.39-2.12) | 1.60 (1.29-2.00) |
|  | **2** | 1.90 (1.40-2.54) | 1.92 (1.49-2.54) | 1.95 (1.57-2.46) | 1.99 (1.57-2.62) |
|  | **3** | 2.18 (1.81-2.85) | 2.21 (1.74-2.63) | 2.24 (1.84-2.71) | 2.15 (1.76-2.54) |

All analysis was conducted using the R language (version 3.6.3). We adopted and modified the code provided by Kucharski et al. (*29*). For reading and manipulating data we used readxl, magrittr, tidyverse, tidyr,DMwr and plyr R packages in addition to pre-installed packages. For processing dates we used R package lubridate. We leveraged R package ggplot2, ggpubr, sf and RColorBrewer for plotting different figures including maps. R package factoextra was used for visualizing PCA. For parallelization, we used for each, doMC and mgcv. All data and code for simulation are available at the following link: https://github.com/rizvi23061998/estimate_chinese_r0

## RESULTS

As of April 7, 2020, total cases reported in China were 83,845 (including Wuhan). The average of daily mean temperature ranged from −25.0560 °C to 21.3948 °C.

### Three regions in different clustering

All four clustering (Table 1) produced similar regions (Figure 1) and predicted similar R_0_ values (Table 2 & Figure 2). In particular, the prefectures are in fact geographically grouped into regions automatically despite that, no direct geographical features have been used for clustering (Table 1 & Supplement Table 1). From the PCA analysis (Figure 2 supplement), it is evident that temperature profile played a significant role, and it is easily noticeable that the increase in temperature increases the R_0_ value.

An interesting point can be noticed in Clustering D (Figure 1(d)) as follows. It exhibits two light green (medium R_0_) prefectures (namely, Tangshan and Binzhou) in the eastern region, which is otherwise assigned green (low R_0_) in other clustering. The fact that Clustering D considers population profile in more detail than other clustering, has played a role here. As it turns out, the ratio of aged population is markedly higher in these two prefectures which essentially put these into the immediate higher R_0_ region. In the same context, we also notice another apparently anomalous prefecture (i.e., Turfan) at the north-western region, which is light green (medium R_0_) in Clustering D as opposed to being green (low R_0_) in other clustering A and C. The analysis shows that the aged population group is not prominent in Turfan; however, further analysis reveals that the sufficiently lower temperature therein has played its part and overshadowed the effect of the aged population. Thus, it seems that we can give higher rank to temperatures in terms of contribution towards transmission than population age.

### Regional Transmission Levels

Region 3 has been identified to be the highest transmission region (among the three) according to all four clustering and the predicted R_0_ for all clustering remains at around 2.19 (2.18 ~ 2.24). The low (medium) transmission region is Region 1 (*2*). In both cases, the predicted R_0_ for all clustering are quite similar, at around 1.62 (1.58 ~ 1.70) for Region 1 and around 1.94 (1.90 ~ 1.99) for Region 2.

Notably, due to the stochastic property of the Monte Carlo simulation, we can safely assume this range as acceptable (Figure 3).

## DISCUSSION

This is the first study to determine the impact of temperature and other potential risk factors through a clustering exercise and to estimate the revised R_0_ of COVID-19 in different regions in China. We have applied the clustering algorithms using different combination of relevant features to ensure a confident and coherent analysis. Our results strongly indicated that R_0_ may be affected (i.e., worsen) by higher temperature, and the prefectures having older population likely favoured its transmission.

Clustering E is based on temperature profile only and it clearly suggested that higher temperature produced higher R_0_. This is a unique finding in itself as the role of temperature in the spread of COVID-19 has been studied (*15-22*) and the predominant prediction was mostly the opposite; evidence from published studies documented negative associations between increasing temperature and COVID-19 transmission (*15, 17, 22*).

Also, population turned out to be an important discriminant feature in clustering, and from the results it can be inferred that higher fraction of aged population is a risk factor and contributes to higher R_0_. While identifying region 3, temperature was the most contributing factor following the demographic factor. Region 3 (exhibiting the highest R_0_) is the warm region where average of the daily mean temperature is around 8.57 ^0^ C whereas Region 1 and Region 2 has this value below 0^0^ C.

We estimated that the mean R_0_ for the COVID-19 ranges from 1.58 to 2.24 (Table 2) and is significantly larger than 1 and is consistent with the WHO’s estimation for the human-to-human (direct) transmission ranged from 1.4 to 2.5 (*33*). On the other hand, most of the predicted mean R_0_ ranging from 2 to 5, and is largely inconsistent with our results (*11*). The reason might be that active surveillance, contact tracing, quarantine, and early strong social distancing efforts contributed to stop transmission of the virus and significantly decreased the effective reproduction number of COVID-19 in China (*10*). Published studies also suggest that the first lockdown resulted in a 65% decrease of the reproduction factor R_0_ and the second, stricter wave of measures eventually managed to bring it to close to 0 (*34*).

Several factors can influence COVID-19 transmission, including environmental variables, population density, and strong public health infrastructures (*35*). The previous studies have seldom analyzed the effects of temperature on the development of COVID-19 in a large scale. However, the association between COVID-19 and temperature was not consistent and changes in temperature showed no significant correlation with cases transmitted, deaths or recovered (*16*). Our findings at the prefecture level on the impact of temperature conditions over the transmission of COVID-19 are not consistent with other published studies (*35, 36*). However, our results are consistent with Luo et al., 2020 (*37*), which reported that the changes in temperature as spring and summer months might not lead to decline of confirmed case counts without the implementation of extensive public health interventions.

The longer the study period, the more stable the model results are expected to be. Temperature may even have no role in transmission since winter is over and temperature started increasing and cases are increasing in warm areas. Control effort, health behavior, social distance has a very big role in R_0_ estimation and can inform public health officials and decision makers about where to improve the allocation of resources, testing sites; also, where to implement stricter quarantines and travel bans (*38*).

The most important environmental implication proved temperature is a critical factor for COVID-19 transmission, which also deserve to be better studied in other regions during this pandemic (*17*).

This study has several limitations, most notably the fact that these analyses were based on routinely collected data during pandemic with the potential for both over and under reporting of COVID-19 cases. Secular changes in reporting could have biased incidence estimates and errors could have been introduced as data were aggregated at higher levels of the health information system. Data accuracy and completeness were not systematically assessed. Some cases might be detected based on clinical signs and symptoms, with the potential for misclassification (*39*). Testing systems have also limited sensitivity and specificity and are particularly likely to misclassify individuals. Parts of China were forced to shut down with restrictions on the population movement. These intervention measures are expected to also have an important impact on the associations between meteorological factors and the transmission of the virus. Additional climatic factors, socio-economic development, population mobility, population immunity, social distancing, health behavior changes, and urbanization, presumably affected the dynamics of the COVID-19 epidemic in China, but we cannot consider every factor in this study. The observed associations between temperature, demographic factors and estimated Ro were ecologic and not at the level of individuals.

### Conclusion

In conclusions, our study does not support that high temperature can reduce the transmission of COVID-19 and it will be premature to count on warmer weather to control COVID-19 (*20*). A weather with high temperature favors the transmission (*17*). There were spatial heterogeneities in COVID-19 occurrence, which could be attributed to temperature. The reasons for the inconsistency in the impact of meteorological factors on COVID-19 among prefectures need further study. This research provides a novel methodology for the global health authorities. As high temperature is associated with the transmissibility of COVID-19, new policies to reinforce health systems or social isolation methods can be adopted, depending on temperature.

## Data Availability

The datasets used and/or analysed during the current study are available at (https://github.com/rizvi23061998/estimate_chinese_r0)

## SUPPLEMENTARY MATERIALS

## Acknowledgement

The authors thank all labs and fields staffs in China for providing diagnosis, treatment to COVID-19 patients in China.

## Funding

UH was supported by the Research Council of Norway (grant # 281077).

Wenyi Zhang was partly supported by grants from the Chinese Major grant for the Prevention and Control of Infectious Diseases (No.2018ZX10733402-001-004, 2018ZX10713003).

## Author contributions

IMK, MSR, UH: Conceptualization, Methodology, Writing - original draft, Data analysis: IMK, MSR, UH. Data preparation: SJ, WZ, JH, and UH. Writing - review & editing, SJ, WZ, and JH. All authors read and approved the final version of the manuscript.

## Competing interests

None declared.

## Data and materials availability

The datasets used and/or analysed during the current study are available (https://github.com/rizvi23061998/estimate_chinese_r0)

**Fig. 1. Supplement:**
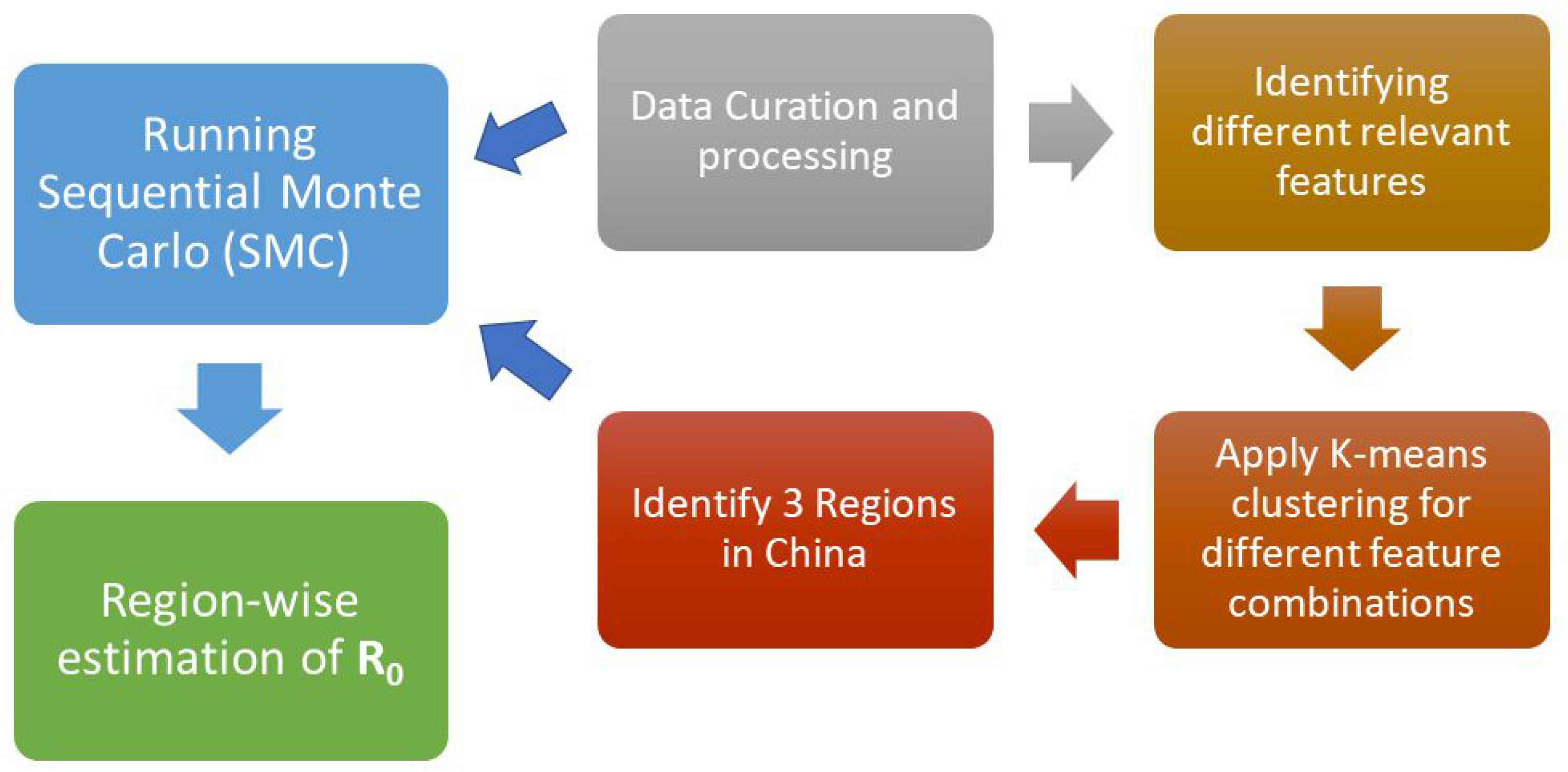
Workflow for revise R0 estimation.

**Fig. 2. Supplement:**
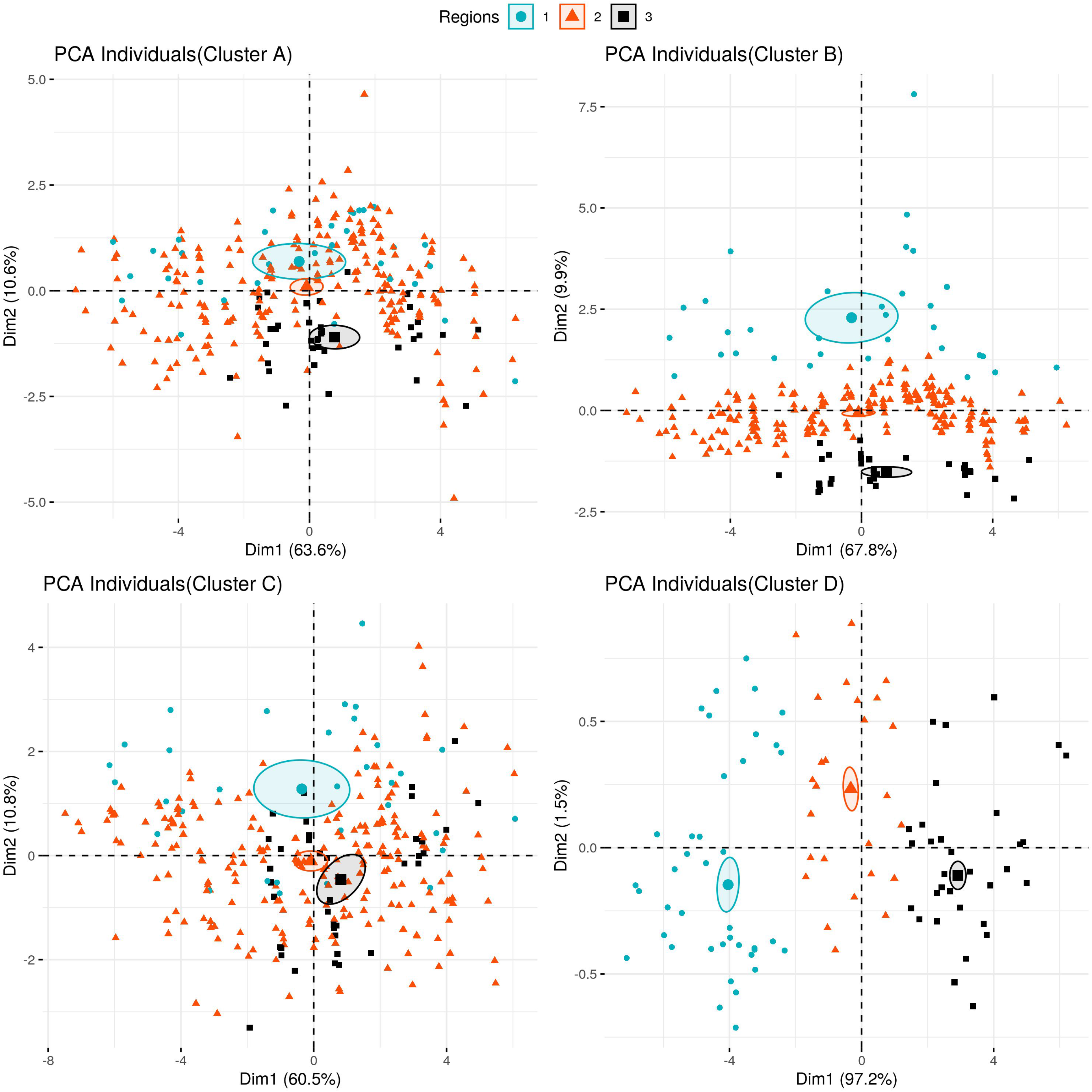
Importance of different features and correlation between them. The length of the arrow refers to the importance of the corresponding feature. Angle between the arrows refers to the correlation between features. The closer two arrows are, the stronger their correlation is.

**Fig. 3. Supplement:**
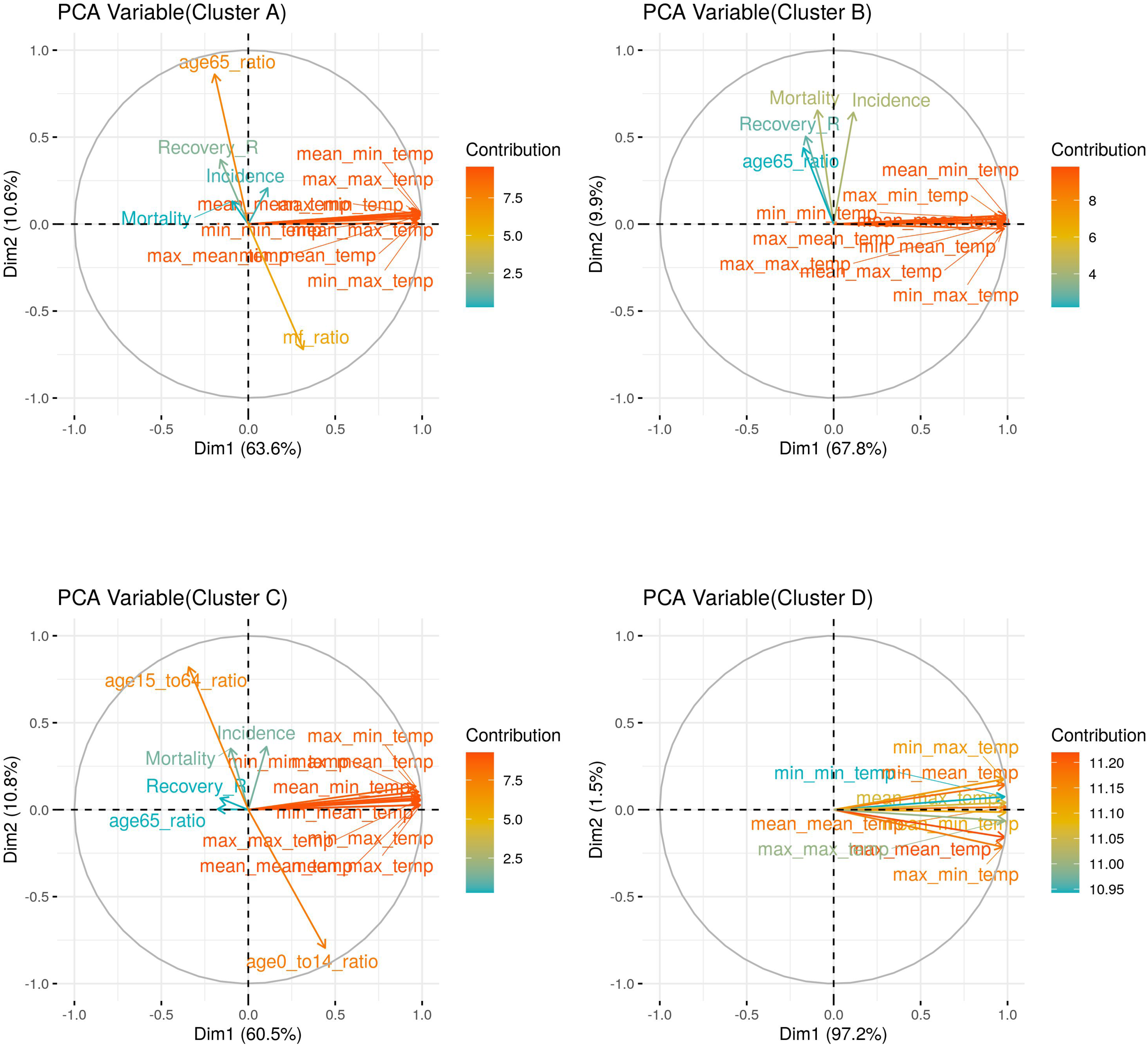
Figures A, B, C, D indicates the clustering based on different sets of features.

---

**Table 1 (Supplement):** List of criteria used by the clustering algorithm

| Criteria/Feature | Comments |
| --- | --- |
| Incidence (I) | ‘I’ is a weak indicator if the incidence is high (Eqn. 1). |
| Recovery Rate (RR) | High ‘RR’ is an indicator of strong health infrastructures (Eqn. 2). |
| Mortality Rate (MR) | High ‘MR’ is a very weak indicator (Eqn. 3). |
| Male/Female Ratio (MFR) | MFR is a significant number of the more male population compared with the female population. If there is a more male population in the prefecture, that will be considered as high risk. |
| Age Group Ratio (AGR) | If any prefecture has a significant number of an older population group, that prefecture should be considered as high risk; AGR has been computed for three groups, namely, $AGR < 15$ (age below 15), $AGR_{15-64}$, (age within the range 15-64) and $AGR > 64$ (age over 64). |
| Temperature (T) | Temperature is speculated to have some effect on transmission. Here, minimum ($T_{min}$), mean ($T_{avg}$), maximum ($T_{max}$) of daily temperatures have been used. |

**Table 2 (Supplement):** RMSE values calculated from the cumulative real cases and cumulative predicted cases. A, B, C, D, refer to the clustering schemes described in Table 1 (supplement). The values within brackets indicate the total number of incidences for that region.

|  |  | **Clustering** |  |  |  |
| --- | --- | --- | --- | --- | --- |
|  |  | **A** | **B** | **C** | **D** |
| *Regions* | 1 | 178 (1341) | 178 (1341) | 190 (1408) | 180 (1341) |
|  | 2 | 1531 (8428) | 1292 (6981) | 1322 (6914) | 2270 (14142) |
|  | 3 | 2829 (18513) | 3046 (19960) | 2913 (19960) | 1966 (12799) |

## Notes

### Competing Interest Statement

The authors have declared no competing interest.

## REFERENCES

1. S. R. Weiss, J. L. Leibowitz, in Advances in virus research. (Elsevier, 2011), vol. 81, pp. 85–164.

2. Y. Yin, R. G. Wunderink, MERS, SARS and other coronaviruses as causes of pneumonia. Respirology 23, 130–137 (2018).

3. N. Zhu et al., A novel coronavirus from patients with pneumonia in China, 2019. New England Journal of Medicine, (2020).

4. .

5. world Health Organization (WHO). (2020), vol. 2020.

6. Q. Li et al., Early Transmission Dynamics in Wuhan, China, of Novel Coronavirus-Infected Pneumonia. N Engl J Med, (2020).

7. C. Huang et al., Clinical features of patients infected with 2019 novel coronavirus in Wuhan, China. The Lancet, (2020).

8. N. C. Peeri et al., The SARS, MERS and novel coronavirus (COVID-19) epidemics, the newest and biggest global health threats: what lessons have we learned? Int J Epidemiol, (2020).

9. D. S. Hui et al., The continuing 2019-nCoV epidemic threat of novel coronaviruses to global health—The latest 2019 novel coronavirus outbreak in Wuhan, China. International Journal of Infectious Diseases 91, 264–266 (2020).

10. S. Sanche et al., High Contagiousness and Rapid Spread of Severe Acute Respiratory Syndrome Coronavirus 2. Emerg Infect Dis 26, (2020).

11. S. Zhao et al., Preliminary estimation of the basic reproduction number of novel coronavirus (2019-nCoV) in China, from 2019 to 2020: A data-driven analysis in the early phase of the outbreak. Int J Infect Dis 92, 214–217 (2020).

12. Jin, JM., Bai, P., He, W., Wu, F., Liu, XF., Han, DM., Liu, S., Yang, PK. Gender differences in patients with COVID-19: Focus on severity and mortality. Front. Public Health, 29 April 2020.

13. C. C.-R. Team, Geographic Differences in COVID-19 Cases, Deaths, and Incidence - United States, February 12-April 7, 2020. MMWR Morb Mortal Wkly Rep 69, 465–471 (2020).

14. Zhang, Y., Jiang, B., Yuan, J., Tao, Y. (2020). The impact of social distancing and epicenter lockdown on the COVID-19 epidemic in mainland China: A data-driven SEIQR model study. Available at [https://www.medrxiv.org/content/10.1101/2020.03.04.20031187v1.full.pdf], last accessed 05.08.2020.

15. A. Tobias, T. Molina, Is temperature reducing the transmission of COVID-19? Environ Res 186, 109553 (2020).

16. Pawar, S., Stanam, A., Chaudhari, M., Rayudu, D. Effects of temperature on COVID-19 transmission. Available at [https://www.medrxiv.org/content/10.1101/2020.03.29.20044461v1], last accessed 05.08.2020.

17. J. Liu et al., Impact of meteorological factors on the COVID-19 transmission: A multi-city study in China. Sci Total Environ 726, 138513 (2020).

18. H. Qi et al., COVID-19 transmission in Mainland China is associated with temperature and humidity: A time-series analysis. Sci Total Environ 728, 138778 (2020).

19. Z. Sun, K. Thilakavathy, S. S. Kumar, G. He, S. V. Liu, Potential Factors Influencing Repeated SARS Outbreaks in China. Int J Environ Res Public Health 17, (2020).

20. Y. Yao et al., No Association of COVID-19 transmission with temperature or UV radiation in Chinese cities. Eur Respir J, (2020).

21. M. F. F. Sobral, G. B. Duarte, A. I. G. da Penha Sobral, M. L. M. Marinho, A. de Souza Melo, Association between climate variables and global transmission oF SARS-CoV-2. Sci Total Environ 729, 138997 (2020).

22. P. Shi et al., Impact of temperature on the dynamics of the COVID-19 outbreak in China. Sci Total Environ 728, 138890 (2020).

23. T. Novel Coronavirus Pneumonia Emergency Response Epidemiology, [The epidemiological characteristics of an outbreak of 2019 novel coronavirus diseases (COVID-19) in China]. Zhonghua Liu Xing Bing Xue Za Zhi 41, 145–151 (2020).

24. Lloyd, S. P. (1957). Least squares quantization in PCM. Technical Report RR-5497, Bell Lab.

25. MacQueen, J. (1967). Some methods for classification and analysis of multivariate observations. Proceedings of the fifth Berkeley symposium on mathematical statistics and probability (pp. 281–297). Oakland, CA, USA.

26. Hartigan, J. A., Wong, M.A. Algorithm AS 136: A K-Means Clustering Algorithm. Journal of the Royal Statistical Society. Series C (Applied Statistics), Vol. 28, No. 1 (1979), pp. 100–108.

27. Q. Li et al., Early Transmission Dynamics in Wuhan, China, of Novel Coronavirus-Infected Pneumonia. N Engl J Med 382, 1199–1207 (2020).

28. Kermack, WO., McKendrick, AG. (1927). A contribution to the mathematical theory of epidemics. Proceedings of the Royal Society of London. Series A, Containing Papers of a Mathematical and Physical Character, 700–721.

29. A. J. Kucharski et al., Early dynamics of transmission and control of COVID-19: a mathematical modelling study. Lancet Infect Dis 20, 553–558 (2020).

30. Jodrá, P. (2012). Computing the asymptotic expansion of the median of the erlang distribution. Mathematical Modelling and Analysis, 17(2), 281–292.

31. Kraemer, M. (2020). Epidemiological data from the nCoV-2019 outbreak: early descriptions from publicly available data. Available at [http://virological.org/t/epidemiological-data-from-the-ncov-2019-outbreak-early-descriptions-from-publicly-available-data/337], last accessed 05.08.2020.

32. Liu, T., Hu, J., Xiao, J., He, G., Kang, M., Rong, Z., Lin, L., Zhong, H., Huang, Q., Deng, A., Zeng, W., Tan, X., Zeng, S., Zhu, Z., Li, J., Gong, D., Wan, D., Chen, S., Guo, L., Li, Y., Sun, L., Liang, W., Song, T., He, J., Ma, W. (2020). Time-varying transmission dynamics of Novel Coronavirus Pneumonia in China. Available at [https://www.biorxiv.org/content/10.1101/2020.01.25.919787v2], last accessed 05.08.2020.

33. World Health Organization Laboratory testing for 2019 novel coronavirus (2019-nCoV) in suspected human cases, World Health Organization (WHO). 2020. Available at [https://www.who.int/health-topics/coronavirus/laboratory-diagnostics-for-novel-coronavirus], last accessed 05.12.2020.

34. K. Prem et al., The effect of control strategies to reduce social mixing on outcomes of the COVID-19 epidemic in Wuhan, China: a modelling study. Lancet Public Health 5, e261-e270 (2020).

35. Wang, M., Jiang, A., Gong, L., Luo, L., Guo, W. (2020) Temperature significant change COVID-19 Transmission in 429 cities. Available at [https://www.medrxiv.org/content/10.1101/2020.02.22.20025791v1], last accessed 05.12.2020.

36. D. N. Prata, W. Rodrigues, P. H. Bermejo, Temperature significantly changes COVID-19 transmission in (sub)tropical cities of Brazil. Sci Total Environ 729, 138862 (2020).

37. Luo et al., 2020. The role of absolute humidity on transmission rates of the COVID-19 outbreak medRxiv (2020), Article 2020.2002.2012.20022467, 10.1101/2020.02.12.20022467.

38. Battiston, P., Gamba, S. (2020). COVID-19: R_0_ is lower where outbreak is larger. Available at [https://arxiv.org/pdf/2004.07827.pdf], last accessed 05.13.2020.

39. T. K. Tsang et al., Effect of changing case definitions for COVID-19 on the epidemic curve and transmission parameters in mainland China: a modelling study. Lancet Public Health 5, e289-e296 (2020).

